# Continuous Aerosol Medication Therapy in an *in vitro* High-Flow System Using Wire Mesh Technology

**DOI:** 10.1101/2020.02.10.20020719

**Authors:** Jhaymie Cappiello, Carla Bremenour, Jason Boyle, Jessica Lumbard, Kamrouz Ghadimi

## Abstract

**BACKGROUND:** Jet nebulizers are commonly used to provide continuous aerosolized medication therapy (CAMT). We observed the function of our CAMT system that utilizes the Aeroneb Solo nebulizer system (Aerogen Ltd, Galway, Ireland). METHODS: An observational study was performed on 2 CAMT systems with 15 Aeroneb nebulizers for each system. CAMT was simulated for 1, 2 and 3 hours. Continuous nebulization was monitored and residual volumes were recorded at the end of each simulation. Our primary endpoint was established as intermittent nebulization observed by nebulizer filling of > 1 ml during CAMT simulation. Secondary endpoint was a residual volume of < 0.1 ml.

**RESULTS:** Out of 30 simulations in two arms, a fluid level was observed to accumulate intermittently in three nebulizers with a residual volume of 0.7 mls in one of these three. This produced a total success rate of 90%, Arm-A 80%, Arm B-100%, for our primary endpoint. Our secondary endpoint was achieved in 29 of the 30 nebulizers for an overall 97% success rate, Arm A-93%, Arm B-100%. CONCLUSION: Our Aerogen Solo CAMT system successfully emitted the set dose with 90% accuracy.

## Introduction

Continuous aerosolized medication therapy (CAMT) is used by many facilities to deliver albuterol and epoprostenol. This therapy is most often provided with mechanical ventilation (MV), noninvasive ventilation (NIV) and through high-flow high-humidity devices (HFHH). Historically, a jet nebulizer has been used for CAMT under these circumstances (1,2). Vibrating mesh technology is reported to deliver a higher respirable dose and improved particle deposition than jet nebulizers without additional flow to the patient breathing circuit (3, 4, 5, 6). These benefits make vibrating mesh technology an attractive alternative to the jet nebulizer when providing CAMT. Our facility utilizes this technology for patients receiving MV, NIV, and HFHH that require CAMT.(fig 1) The only clinically available vibrating mesh product for use in this fashion is the Aeroneb Solo (Aerogen, Galway, Ireland). We designed an observational study with a primary endpoint of observed continuous nebulization and a secondary endpoint of a residual volume at the end of testing procedures to be <0.1 mls. The authors have no conflicts of interest to report.

**Figure 1.**
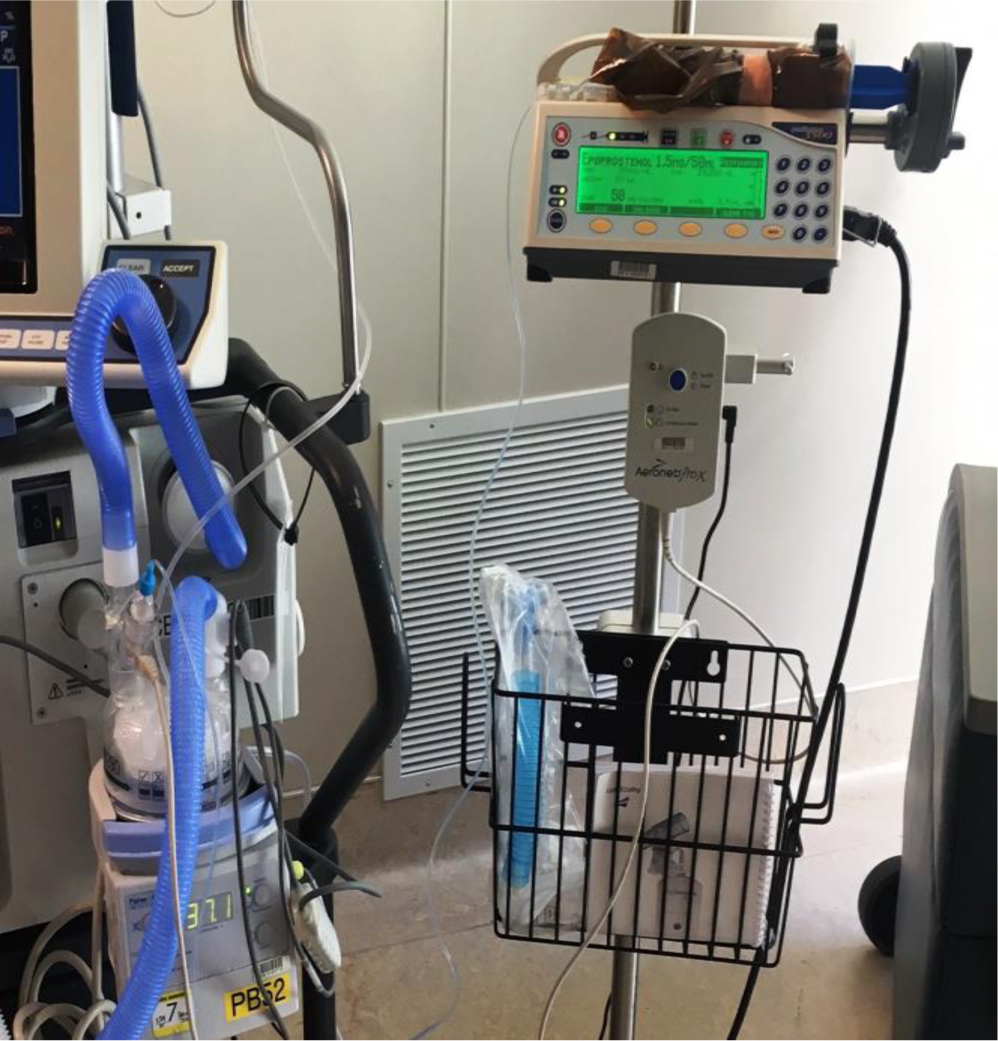

## Methods

We developed a two-armed *in vitro* observational study of our CAMT Aeroneb Delivery System operating under an established testing protocol. Two CAMT units were taken out of our service fleet and labeled A and B. Each arm had identical equipment: an Aerogen PRO-X controller, control module cable, AC adapter, Venturi adapter, Aerogen T piece, and a Medfusion 3500 syringe pump Smiths Medical, Dublin, Ohio). A new Aeroneb Solo was used for each test and the Venturi adapter (Carefusion, Yorba Linda, California) was set to deliver a constant flow of approximately 50 lpm through the Aerogen T piece. Normal saline was used and delivered via the Aerogen Continuous Nebulization Tube Set assembly at a rate of 7.51 ml/hr. The Aeroneb Solo was placed in the Aeroneb Solo T piece and secured in a vertical position. (Fig 2) Each arm consisted of 1 hour, 2 hour, and 3 hour runs. Each run consisted of 5 nebulizers. Prior to each run, the nebulizer was tested with 1 ml of normal saline to ensure function and ran to dry. The Aerogen PRO-X module was set in continuous mode. Upon completion of the run time, the syringe pump was stopped, the nebulizer removed from the “T” piece, and the module controller was turned off. Residual volume was then measured by syringe extraction. The nebulizers were constantly monitored for filling, nebulization and untouched by the study team during the testing. Recorded values included: start/stop times, volume delivered, residual volume, and filling. A nebulizer failure was defined as development of a fluid level > 1ml during nebulization and/or a residual volume of >0.1 ml. Eight Respiratory Care Practitioners participated in performing these tests.

**Figure 2.**
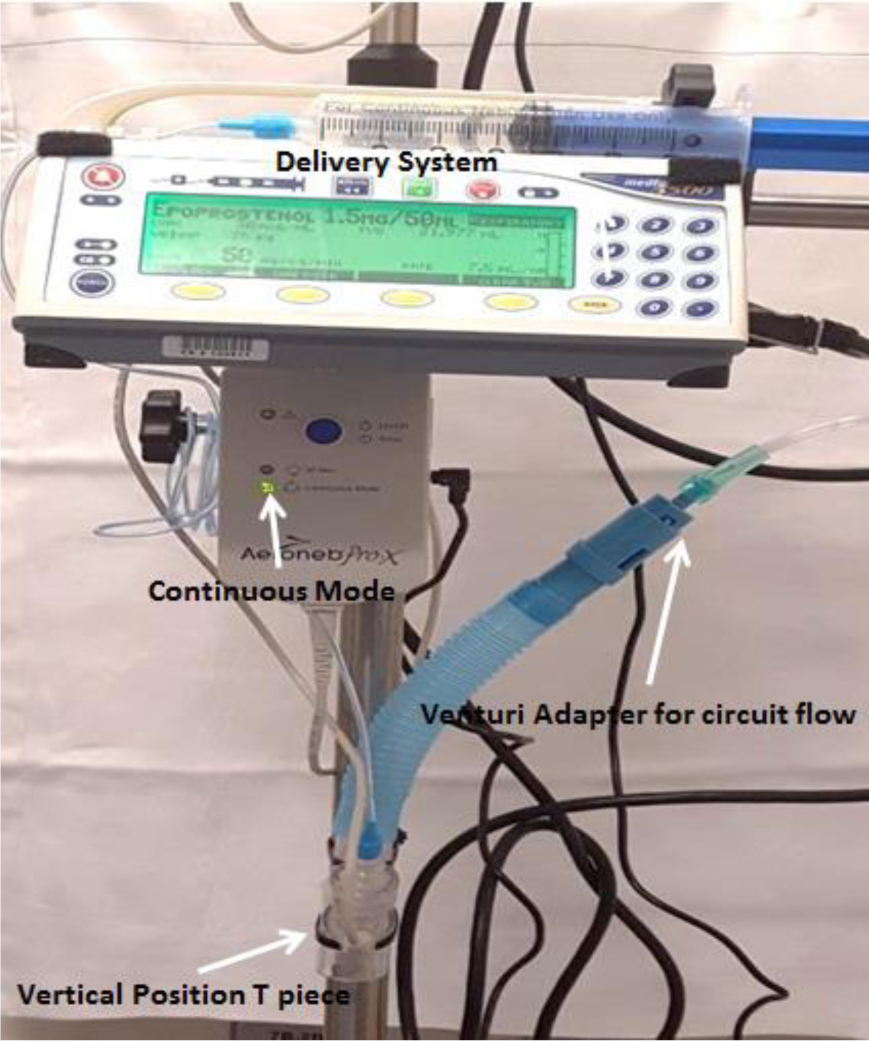

## Results

All nebulizers chosen for testing passed their initial function test. A fluid level was observed to accumulate intermittently in three nebulizers during their observational runs. This produced a total success rate of 90%, Arm-A 80%, Arm B-100%, for our primary endpoint. Our secondary endpoint of a residual volume of <0.1 ml was achieved in 29 of the 30 nebulizers for an overall 97% success rate, arm A-93%, arm B-100%. Table 1 shows the delivered and residual volumes for the observations performed. Delivered volume variance was based on syringe pump shut off time and was not considered impactful on endpoints. Observed residual volume range was <0.1ml – 0.7 mls.

**Table 1.** Delivered and residual volumes according each in vitro high-flow system.

|  | <u>System A - Volumes</u> |  | <u>System B - Volumes</u> |  |
| --- | --- | --- | --- | --- |
|  | §Delivered | §Residual | §Delivered | §Residual |
| 1 hour | 7.51 | <0.1 | 7.41 | <0.1 |
|  | 7.51 | <0.1 | 7.51 | <0.1 |
|  | 7.51 | <0.1 | 7.51 | <0.1 |
|  | 7.51 | <0.1 | 7.48 | <0.1 |
|  | 7.46 | 0.7* | 7.61 | <0.1 |
| 2 hours | 14.98 | <0.1 | 15.06 | <0.1 |
|  | 15.02 | <0.1* | 15.22 | <0.1 |
|  | 15.01 | <0.1 | 15.04 | <0.1 |
|  | 15.02 | <0.1 | 15.16 | <0.1 |
|  | 15.04 | <0.1 | 15.00 | <0.1 |
| 3 hours | 22.05 | <0.1 | 22.6 | <0.1 |
|  | 22.43 | <0.1* | 22.33 | <0.1 |
|  | 22.48 | <0.1 | 22.45 | <0.1 |
|  | 22.50 | <0.1 | 22.54 | <0.1 |
|  | 22.49 | <0.1 | 22.44 | <0.1 |

\*Intermittent nebulization observed
§All volumes measured in milliliters (ml)

## Discussion

Our *in vitro* study was designed to observe the function of our CAMT system that incorporates the Aeroneb Solo system in simulated clinical use. Our definition of failure was based on our expectation that the system will deliver a constant rate of nebulization as the drug was delivered by our high-flow administration system. Before this study, our delivery system was observed to provide a nebulization interval of 42 seconds at a delivery rate of 7.51 ml/hr. By manufacturers’ recommendation, the maximum input rate for the Aeroneb Solo when operating in continuous nebulization mode is 12 ml/hr. If properly functioning, fluid accumulation in the fill chamber within the Aerogen T-piece of >0.1 ml should not be appreciated for any given delivery dose. During our study, the observed rate of fluid accumulation >0.1 ml was 10%. Of note, the three nebulizers that functioned intermittently were in the system “A” arm of the study while no failures were observed in system “B”. This suggests that other components of the delivery system may have been the issue and not necessarily a discrete nebulizer dysfunction. Intermittent nebulization may impact dose response in agents with a short half-life such as epoprostenol (half-life 6 minutes). (7) The residual volume of >0.1 ml (0.7mls) found in one test may also be due to non-nebulizer factors. A potential contributor to this residual volume may be our method of removing the administration tubing from the nebulizer for syringe extraction. Fluid could have been tapped off into the nebulizer as it was removed. This explanation however, is unlikely due to the large number of nebulizers successfully meeting a residual volume of <0.1 ml. The addition of flow in our system may have contributed to our minimal failure rate, which is an important component that has been omitted in a previous study by Gowda et al. (8) In this in vitro study without flow, the authors theorized that the gravitational feed in these devices may be a contributing factor in their observed failure rates. The benefit of the flow concept is best explained by the Bernoulli Principle with the corollary of the Venturi effect and may have contributed to “vacuuming” the generated aerosol from the wire mesh into the gas delivery system. As the gas delivery system forces flow through the airway circuit and past the Aerogen “T”, lateral pressure decreases and draws the aerosol in for delivery, which may have optimized aerosolization. (Fig 3)

**Figure 3.**
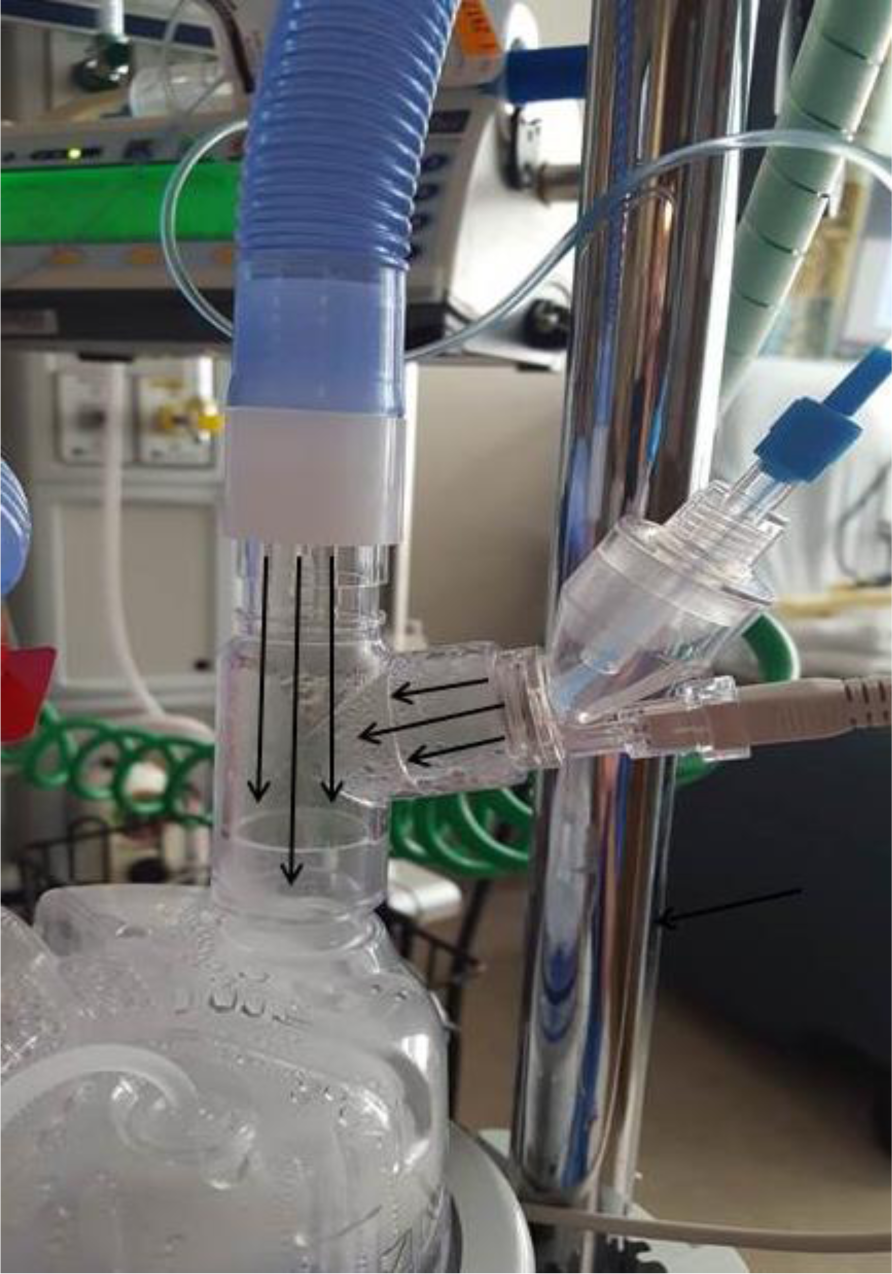

Limitations of our study include our process for measuring residual volume. Weighing the nebulizers’ pre and post would have accounted for any residual volume not obtained with syringe extraction. Our choice of adding continuous flow to the delivery system was to replicate delivery in the clinical setting during CAMT (e.g., MV, NIV, and HFHH). Our facility places these nebulizers on the dry side of the humidifier when applicable and at the outlet port if no humidifier is used. (9, 10, 11) This may result in the nebulizer “T” being in either the vertical or horizontal position. The vertical position was utilized during this *in vitro* investigation. The manufacturer indicates that both are acceptable but the vertical position may have varying delivery volume characteristics than in the horizontal position. Skaria and Smaldone demonstrated product variability in nebulizer output in relation to position. (12) We did not test the failed nebulizers from system A on system B in order to elucidate a factor other than the nebulizer.

This investigation was an observational study of our CAMT Aeroneb Solo nebulizer system set in continuous mode with established time frames only (1, 2, and 3 hour intervals). Failure rates clinically reported may be due to other factors. These factors include but are not limited to: power chord dislodgement triggering the device to enter the 30-minute nebulization mode, control module failure, control module cable dislodgement resulting in cessation of nebulization, control module cable malfunction, and syringe pump malfunction.

## Conclusion

The Aeroneb Solo system, based on our established criteria, successfully emitted the set dose with 90% accuracy. Respiratory Care departments may find this information useful when developing policies regarding CAMT monitoring and equipment maintenance. Incorporation of an audible alarm to elucidate device failure is a correct step in the direction of timely intervention but should not be substituted for vigilance and quality assurance of observed aerosolization and drug delivery. Prospective studies on CAMT with the Aeroneb system are warranted in the high-flow clinical environment in order to determine the clinical significance of this observed nebulization failure.

## Data Availability

Data used is present in the submitted article

